# A ten-year microbiological study of *Pseudomonas aeruginosa* strains revealed diffusion of carbapenems and quaternary ammonium compounds resistant populations

**DOI:** 10.1101/2022.11.06.22282008

**Authors:** Marine Pottier, François Gravey, Sophie Castagnet, Michel Auzou, Langlois Bénédicte, François Guérin, Jean-Christophe Giard, Albertine Léon, Simon Le Hello

**Author notes:** These authors contributed equally to this work and share last authorship (Albertine Léon, Simon Le Hello).

## Abstract

*Pseudomonas aeruginosa* is one of the leading causes of healthcare-associated infections. For this study, the susceptibility profiles to antipseudomonal antibiotics and a quaternary ammonium compound, didecyldimethylammonium chloride (DDAC), widely used as a disinfectant, were established for 180 selected human and environmental hospital strains isolated between 2011 and 2020. Furthermore, a genomic study was performed to determine their resistome and clonal putative relatedness. During the ten-year study period, it was estimated that 9.5% of clinical *P. aeruginosa* were resistant to carbapenem, 11.9% presented an MDR profile, and 0.7% an XDR. Decreased susceptibility (DS) to DDAC was observed for 28.0% of strains that was significantly more associated with MDR and XDR profiles and from hospital environmental samples (p <0.0001). According to genomic analyses, the *P. aeruginosa* population unsusceptible to carbapenems and/or to DDAC was diverse but mainly belonged to top ten high-risk clones described worldwide. The carbapenem resistance appeared mainly due to the production of the VIM-2 carbapenemase (39.3%) and DS to DDAC mediated by MexAB-OprM pump efflux overexpression. This study highlights the diversity of MDR/XDR populations of *P. aeruginosa* which are unsusceptible to molecules that are widely used in medicine and hospital disinfection and are probably distributed in hospitals worldwide.

## Introduction

*Pseudomonas aeruginosa* (*P. aeruginosa*) is a Gram-negative bacillus that is remarkably adaptable and metabolically versatile^[1]^. Ubiquitous in the environment, it can be found in soil, water, and hospital environments such as sinks and drains^[2]^. For humans, *P. aeruginosa* is an opportunistic pathogen responsible for various infections, such as hospital-acquired pneumonia, bloodstream, and urinary tract infections^[3]^. Its presence in humans can be a sign of colonization or chronic or evolving infection^[4]^. *P. aeruginosa* infections occur particularly in critically ill and immunocompromised patients and are largely health care-associated^[5]^.

In France, *P. aeruginosa* was the 4^th^ leading microorganism responsible for nosocomial infections^[6]^ and was shown to increase the total cost of a hospitalization stay^[7]^. On a larger scale, in 2020, *P. aeruginosa* (6.2%) was the 5^th^ most commonly reported bacterial species from invasive isolates (blood or cerebrospinal fluid)^[8]^, and the most frequently isolated microorganism in intensive care unit (ICU)-acquired pneumonia episodes^[9]^ in the European Union/European Economic Area.

One of the challenges of *P. aeruginosa* infections is the difficulty of treatment^[10]^. Due to their intrinsic resistance to many antibiotics, only eight classes of antimicrobials are currently used against *P. aeruginosa* infections: penicillins with β-lactamase inhibitors, carbapenems, monobactams (aztreonam), cephalosporins, aminoglycosides, fluoroquinolones, phosphonic acids (fosfomycin) and polymyxins (colistin and polymyxin B)^[11]^.

Recent drugs such as ceftolozane-tazobactam, cefiderocol, and imipenem-cilastatin/relebactam were developed^[12,13]^. However, *P. aeruginosa* can also acquire some resistance by gene mutational processes, which modify the expression and function of chromosomally encoded mechanisms, or by gene acquisition^[14]^. This problem is such that in 2017, the World Health Organization published a list to prioritize and stimulate research and development of effective drugs, and classified carbapenem-resistant *P. aeruginosa* as a critical priority. Carbapenem resistance constitutes an appropriate marker for multidrug-resistant (MDR), extensively drug-resistant (XDR), and pandrug-resistant bacteria because it usually involves a wide range of co-resistance to unrelated antibiotic classes^[15]^. It is important to note that if not successfully treated in the acute phase, *P. aeruginosa* can establish chronic biofilm infections, which are difficult or even impossible to eradicate^[1,16]^. Because biofilms decrease antimicrobial susceptibility, it is important to diagnose early-stage *P. aeruginosa* infections, keeping in mind the high level of resistance cells can possess even in planktonic mode (MDR/XDR profiles).

Carbapenem resistance can be due to chromosomal mutations that can lead to decreased permeability by the outer membrane protein OprD2 deficiency^[17]^, overexpression of efflux pumps, and hyperproduction of the chromosomal cephalosporinase AmpC or by enzymatic antibiotic modifications^[18,19]^. Finally, acquired carbapenemases among *P. aeruginosa* strains are increasingly reported worldwide, mainly concerned with Ambler class B metallo-β-lactamase (VIM and IMP) or Ambler’s class A (KPC and GES variants) enzymes^[20–22]^. The presence of strains carrying highly transferable carbapenemases represents a major health threat, and it is responsible for outbreaks in wards hosting immunocompromised patients. In addition, the overuse and misuse of disinfectants can also lead to decreased susceptibility to medically important antimicrobials due to cross-resistance and/or co-resistance mechanisms^[23]^. For example, in *P. aeruginosa* antibiotic resistances can be led by exposure to sodium hypochlorite, didecyldimethylammonium chloride (DDAC)^[24]^, or chlorhexidine^[25]^. These can occur by natural selection or by reinforcement of an acquired resistance mechanism that allows for adaptation to the new environment^[24]^.

For care facilities, disinfectants are considered the first option against pathogen dissemination^[26]^. In the European Union, there are approximately 250 different chemical compounds used, either alone or in combination, in disinfectant products including quaternary ammonium-based disinfectants^[27]^. DDAC is a compound used as a biocide in various applications such as agricultural, food, leisure, and medical equipment, both for private and professional use^[28]^. In health products, it is used for its detergent and disinfectant actions on floors, walls, accessories, examination tables, medicated equipment, and noninvasive medical devices^[29]^.

The present study focused on *P. aeruginosa*, both clinical and environmental strains, isolated in a French University Hospital over ten years, 2011-2020, to 1) describe the circulating populations, their genetic diversity, their antimicrobial resistance profiles and resistomes, 2) determine their susceptibility to the DDAC detergent disinfectant, and 3) look for potential epidemiological and resistance mechanisms links.

## Results

### Antimicrobial and DDAC resistance phenotypes

Regarding the AST data available, 4,375 of 6,661 P strains were tested for all 8 antibiotic classes, 11.9% displayed an MDR profile, 0.7% displayed XDR, and no strains were categorized as pandrug-resistant (Figure 1a and b). The proportion of resistance to carbapenems was evaluated at 9.5% (n=415), with a variable annual frequency from 0.05 [95% CI%: 0.03-0.09] in 2013 to 0.17 [95% CI%: 0.10-0.28] in 2012 and an average of 0.12 [95% CI%: 0.10-0.08] (Figure 2a). Colistin (1.7%), ceftolozane-tazobactam (3.9%) and amikacin (4.6%) were the most active antibiotics (>95%) on clinical *P. aeruginosa* isolated in our university hospital between 2011 and 2020 (Figure 1b). Surprisingly, no variation in the overall level of resistance was observed between the P and H strains.

**FIGURE 1.**
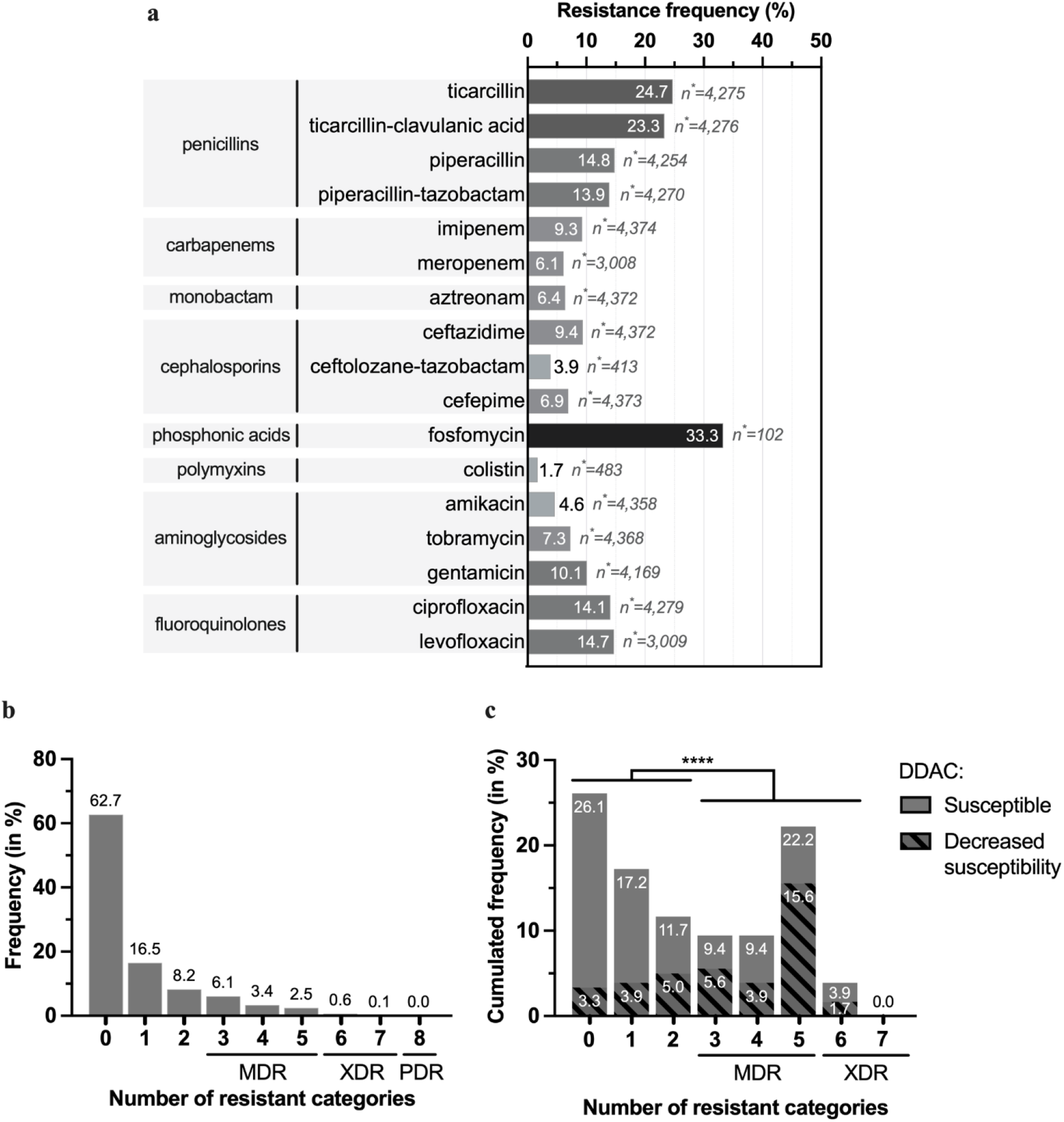
Antimicrobial resistance for hospital strains (n=4,375) and study panel (n=180). (**a**) Occurrence of antibiotics resistance for all *P. aeruginosa* P strains tested for antibiotic susceptibility at the University Hospital of Caen (n=4,375). Number of strains (in percent) showing resistance for 0 to 7 antibiotic classes; for (**b**) all *P. aeruginosa* P strains tested among the total (n=4,375) and (**c**) the study panel (n=180). (**c**) The hatched areas represent profiles that also accumulate DS to the disinfectant DDAC. The different antibiotic categories are penicillins (ticarcillin, ticarcillin-clavulanic acid, piperacillin, piperacillin-tazobactam), carbapenems (imipenem, meropenem), monobactam (aztreonam), cephalosporins (ceftazidime, ceftolozane-tazobactam, cefepime), fosfomycin, aminoglycosides (amikacin, tobramycin, gentamicin) and fluoroquinolones (ciprofloxacin and levofloxacin). Fisher’s test for populations independence for DS to DDAC and loss of susceptibility to more than three categories of antibiotics. n*: number of strains tested for the antibiotic; ***: p-value ≤ 0.0001 after analysis by Fisher’s independence test. DDAC: didecyldimethylammonium chloride; DS: decreased susceptibility; MDR: multidrug-resistant; P: strains isolated from patients; XDR: extensively drug-resistant.

**FIGURE 2.**
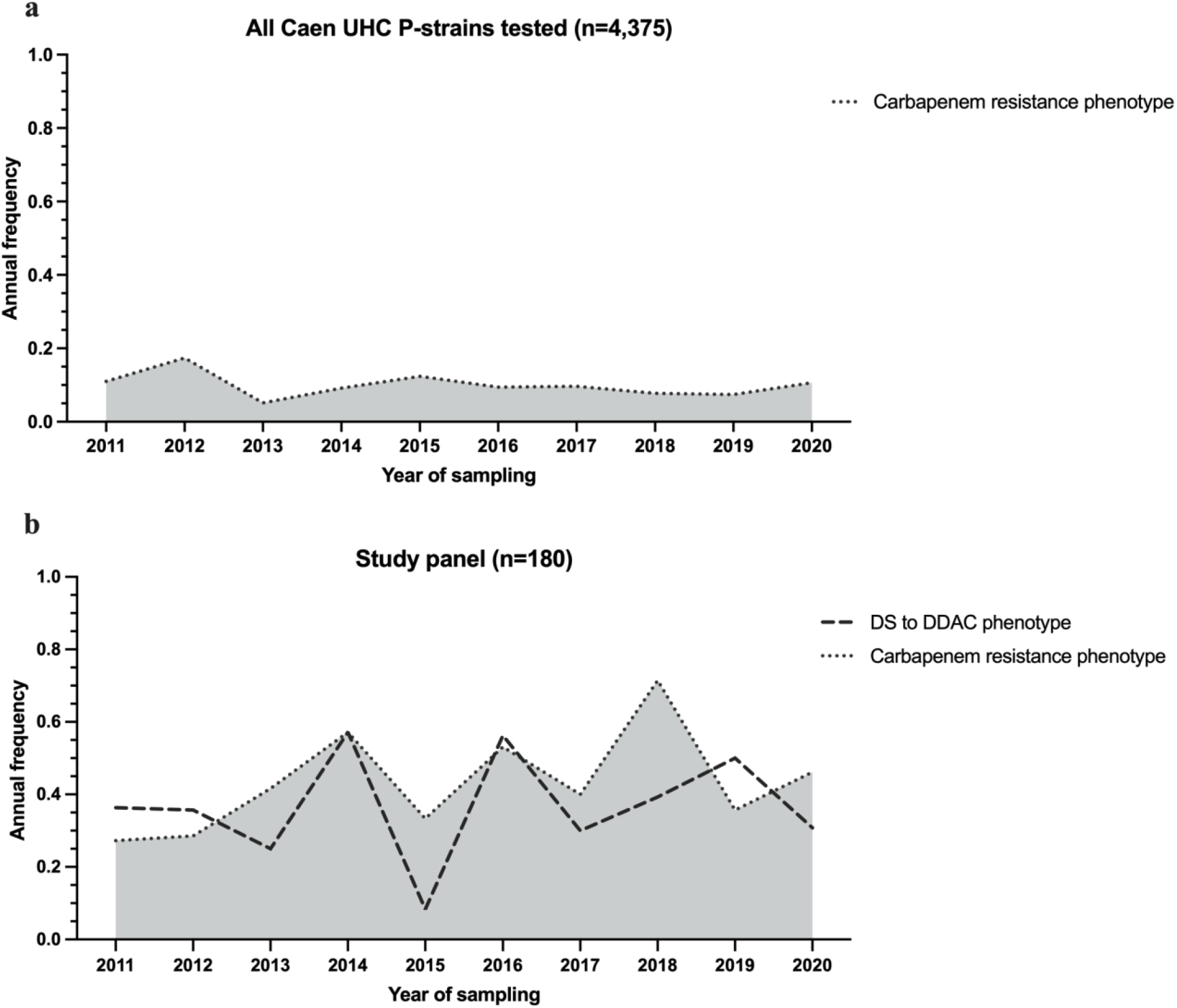
Annual frequency of carbapenem resistance phenotype for all *P. aeruginosa* P strains tested for antibiotic susceptibility at the University Hospital of Caen n=4,375 (a) and the study panel n=180 (b); and annual frequency of the decreased susceptibility to DDAC phenotype on the study panel (b). Strains considered resistant to carbapenems were resistant to imipenem and/or meropenem. DDAC: didecyldimethylammonium chloride; DS: decreased susceptibility.

Among the 180 selected strains of the study panel, all the phenotype of resistance has been selected with 26.1% (n=47) of the strains displaying susceptibility to all 16 antibiotics tested (7 classes), 28.9% (n=52) showing nonsusceptibility to one or two antibiotic classes, 41.1% (n=74) were considered MDR, and 3.9% (n=7) as XDR. (Figure 1c and SD 6a). Carbapenem resistance was 46.7% (n=84/180) of the strains, with an average annual expression frequency of 0.43 [95% CI: 0.34-0.53] (Figure 2b). They were isolated from 13 different sample types, such as sink traps (n=31/51), urine (n=17/24), bronchial aspiration (n=10/16), abscesses (n=6/11), stools (n=4/8), sputum (n=4/11), and blood cultures (n=4/16), from various wards, mainly the ICU (n=34/55) and hematology (n=10/31). Among the carbapenem-resistant *P. aeruginosa* strains, 33 (39.3%) expressed the metallo-β-lactamase VIM-2 enzyme. These last strains were isolated from sink traps (n=15/51), urine (n=6/24), bronchial aspiration (n=5/16), stool (n=4/8), sputum, blood culture, and catheters (1 for each), and were mainly found in the ICU (n=21/55) and hematology (n=4/31) wards. GES-5 was the second most frequent carbapenemase found in strains, with eight cases (SD 1 Part 1).

Regarding the quaternary ammonium antimicrobial (DDAC), MICs between 8 mg/L and 128 mg/L were found, and decreased susceptibility (MIC>62.9 mg/L) was observed in 38.9% of strains (n=70/180) (SD 6a). Statistically, the DS to DDAC phenotype was associated with carbapenem resistance (Fisher’s exact test p value <0.0001). More generally, DS to DDAC association increased with the number of drug-resistant strains: for 3.3% (n=6/180) of strains susceptible to all 7 antibiotic classes tested, for 8.9% (n=16/180) nonsusceptible to one or two categories of antibiotics, for 25.0% (n=45/180) and 1.7% (n=3/180) of strains with MDR and XDR profiles, respectively (Figure 1c). Even if some strains with DS to DDAC had a low antibioresistance level, overall, the DS to DDAC was significantly associated (Fisher’s exact test p value <0.0001) with a high level of antibiotic resistance (nonsusceptibility to more than three antibiotic classes) (Figure 1c). Furthermore, DS to DDAC was found more often in environmental samples (n=35, 62.5% of H strains) than in human samples (n=35, 28.2% of P strains) Fisher’s exact test p value <0.0001 (SD 6b). The phenotype was retrieved each year during the 2011-2020 period. The annual expression frequency variation is shown in Figure 2b. To assess the emergence of *P. aeruginosa* strains with DS to DDAC among clinical strains over the last decade, the random inclusion of 10 strains per year confirmed the high frequency, with 28.0% of strains showing DS to DDAC with an MIC range of 64 to 128 mg/L, mainly found in respiratory samples (n=8/25; 32.0%), surgical pus (n=4/13; 30.8%), urinary samples (n=6/21; 28.6%) and digestive samples (n=4/14; 28.6%).

### Diversity of clinical strains of the study panel and patient information

Among all clinical *P. aeruginosa* strains (n=6,661) isolated over the 2011-2020 period, the 124 study panel strains were isolated from 21 different wards, mainly from ICU (n=34, 27.4%), surgery (n=21, 16.9%), otorhinolaryngology (n=9, 7.3%), pneumology (n=9, 7.3%). Among all environmental *P. aeruginosa* strains (n=6,388), the 56 H strains were mainly from hematology (n=23, 41.1%), ICU (n=21, 37.5%), hepato-gastroenterology (n=6, 10.7%), and cardiology (n=5, 8.9%). Most of clinical P strains were isolated from urine (n=24, 19.4%), bronchial aspiration (n=16, 12.9%), blood culture (n=15, 12.1%), sputum (n=11, 8.9%), and abscess (n=11, 8.9%). Sink trap samples represented 91.1% of H strains (SD 5).

Study panel P strains were isolated from patients aged one month to 97 years old, of which 63.7% (n=79/124) were aged 50 years or older, and most strains were isolated from men (n=81/124, 65.3%). Most of the patients presented altered health status with one or more comorbidities (n=103/118, 87.3%) and/or immunosuppression (n=60/116, 51.7%). Concerning their Charlson Comorbidity Index score, more than 61.5% (n=72/117) had a score ≥3, including 12.0% (n=14) who even had a score ≥7. Most of the strains, 81.9% (n=95/116), were hospital-acquired, and 61.8% (n=76/123) caused infections during the stay. In total 24 patients died, and the cause of death was associated with *P. aeruginosa* for 8.0% (n=9/112) of all patients (excluding non-found cases), most of which were hospital-acquired. In total, 60.8% (n=62/102) of the patients received antipseudomonal treatment after the species identification of the strain (Table 1). Untreated patients were associated with various samples including urine (n=6), sputum (n=3) and catheters (n=3). The most prescribed treatments were piperacillin/tazobactam (n=18, 29.0% of antipseudomonal-treated patients), followed by amikacin (n=16, 25.8%), ceftazidime (n=15, 24.2%), ciprofloxacin (n=15, 24.2%) and imipenem (n=11, 17.7%). Meropenem was only prescribed for 4.8% (n=3) of cases (Table 2).

**TABLE 1.** Demographic and clinical characteristics of patients associated with the P-strains (n=124) from the study panel. Distribution by the population age, sex, presence of comorbidities and patient immune status at the sampling time, the score of the Charlson Comorbidity Index score at the sampling time, type of acquisition and carriage of the strain at the sampling time (evolution of status not considered), whether death was as associated with the presence of *P. aeruginosa*, and whether the patient received an antipseudomonal antibiotic during the stay. * Not found cases were excluded from the total.

|  | Demographic/ clinical variable | Number of patients | % |
| --- | --- | --- | --- |
| Age | <20 | 11 /124 | 8.9 |
|  | 20-29 | 10 /124 | 8.1 |
|  | 30-39 | 13 /124 | 10.5 |
|  | 40-49 | 11 /124 | 8.9 |
|  | 50-59 | 17 /124 | 13.7 |
|  | 60-69 | 22 /124 | 17.7 |
|  | 70-79 | 22 /124 | 17.7 |
|  | 80-89 | 14 /124 | 11.3 |
|  | ≥90 | 4 /124 | 3.2 |
| Gender | M | 81 /124 | 65.3 |
|  | F | 43 /124 | 34.7 |
| Comorbidities and immune status* | Comorbidities | 103 /118 | 87.3 |
|  | Immunocompromised | 60 /116 | 51.7 |
| Charlson Comorbidity Index Score* | 0 | 18 /117 | 15.4 |
|  | 1 | 11 /117 | 9.4 |
|  | 2 | 16 /117 | 13.7 |
|  | 3 | 12 /117 | 10.3 |
|  | 4 | 20 /117 | 17.1 |
|  | 5 | 15 /117 | 12.8 |
|  | 6 | 11 /117 | 9.4 |
|  | ≥7 | 14 /117 | 12.0 |
| Acquisition type and infection/ colonization* | Hospital acquisition | 95 /116 | 81.9 |
|  | Community acquisition | 21 /116 | 18.1 |
|  | Infection | 76 /123 | 61.8 |
|  | Colonization | 47 /123 | 38.2 |
| Deceased* | Death associated with *P. aeruginosa* | 9 /112 | 8.0 |
| Treatment with an antipseudomonal* | Treated before strain identification | 25 /100 | 25.0 |
|  | Treated after strain identification | 62 /102 | 60.8 |

**TABLE 2.** Antipseudomonal antibiotics received by patients associated with the P-strains (n=124) from the study panel, before and after the identification of *P. aeruginosa* during the stay.

| Family | Antibiotic or association | Number of patients with antipseudomonal treatment |  |  |  |
| --- | --- | --- | --- | --- | --- |
|  |  | Before *P. aeruginosa* identification (n=25) |  | After *P. aeruginosa* identification (n=62) |  |
|  |  | n | % | n | % |
| Carboxypenicillins | ticarcillin/clavulanic acid | - | - | 1 | 1.6 |
| Ureidopenicillins | piperacillin | - | - | 3 | 4.8 |
|  | piperacillin/tazobactam | 9 | 36.0 | 18 | 29.0 |
| Carbapenems | imipenem/cilastatin | 3 | 12.0 | 11 | 17.7 |
|  | meropenem | 1 | 4.0 | 3 | 4.8 |
| Monobactams | aztreonam | - | - | 5 | 8.1 |
| Third generation cephalosporins | ceftazidime | 5 | 20.0 | 15 | 24.2 |
| Fourth generation cephalosporins | cefepime | - | - | 4 | 6.5 |
| Phosphonic acid | fosfomycin | - | - | 3 | 4.8 |
| Polymyxins | colistin | 3 | 12.0 | 7 | 11.3 |
|  | polymyxin B | - | - | 1 | 1.6 |
| Aminosides | amikacin | 5 | 20.0 | 16 | 25.8 |
|  | gentamicin | 6 | 24.0 | 2 | 3.2 |
|  | tobramycin | 2 | 8.0 | 5 | 8.1 |
| Fluoroquinolones | ciprofloxacin | 7 | 28.0 | 15 | 24.2 |
|  | levofloxacin | - | - | 2 | 3.2 |

### Genomic diversity and resistome analysis

The 77 sequenced strains of the panel were distributed into ten serotypes (Table 3). Three major serotypes were found for the P strains (O6 at 35.1%, O11 at 21.6%, and O12 at 13.5%) and H strains (O11 at 42.5%, O6 at 22.5%, and O12 at 10.0%). Other serotypes each represented less than 10% of the population.

**TABLE 3.** Serotyping and multilocus sequence typing (MLST) of all sequenced strains: the panel for genomic characterization (n=77) and reference strains (n=4). Each value represents the number of strains for each serotype or sequence type and the associated frequency (in percent). A black cross (x) marks the sequence types considered as the global top 10 *P. aeruginosa* high-risk clones based on prevalence, global spread, and association with MDR/XDR profiles and regarding extended-spectrum β-lactamases and carbapenemases^[32]^. H: strains isolated from the hospital environment; MLST: Multi locus sequence typing; MDR: Multidrug-resistant; P: strains isolated from patients; VIM-2: strains producing the class B carbapenemase VIM-2; XDR: Extensively drug resistant.

| Serotype | Sequence type | High-risk clones | P strains |  | H strains |  | Total |  |
| --- | --- | --- | --- | --- | --- | --- | --- | --- |
|  |  |  | Strain number | % | Strain number | % | Strain number | % |
| **O11** | 298 | X | 2 | 5 | 6 | 15 | 8 | 10 |
|  | 235 | X | 5 | 14 | 2 | 5 | 7 | 9 |
|  | 309 |  |  | 0 | 4 | 10 | 4 | 5 |
|  | 308 | X | 1 | 3 | 3 | 8 | 4 | 5 |
|  | 316 |  |  | 0 | 2 | 5 | 2 | 3 |
| Total O1 |  |  | 8 | 22 | 17 | 43 | 25 | 32 |
| **O6** | 233 | X | 10 | 27 | 6 | 15 | 16 | 21 |
|  | 395 |  | 3 | 8 | 3 | 8 | 6 | 8 |
| Total O6 |  |  | 13 | 35 | 9 | 23 | 22 | 29 |
| **O12** | 111 | X | 5 | 14 | 4 | 10 | 9 | 12 |
| Total O12 |  |  | 5 | 14 | 4 | 10 | 9 | 12 |
| **O4** | 175 | X | 3 | 8 |  | 0 | 3 | 4 |
|  | 111 | X |  | 0 | 1 | 3 | 1 | 1 |
|  | 2407 |  |  | 0 | 1 | 3 | 1 | 1 |
|  | 389 |  |  | 0 | 1 | 3 | 1 | 1 |
| Total O4 |  |  | 3 | 8 | 3 | 8 | 6 | 8 |
| **O10** | 253 |  | 2 | 5 | 3 | 8 | 5 | 6 |
| Total O10 |  |  | 2 | 5 | 3 | 8 | 5 | 6 |
| **O1** | 27 |  | 1 | 3 | 1 | 3 | 2 | 3 |
|  | 207 |  |  | 0 | 1 | 3 | 1 | 1 |
| Total O1 |  |  | 1 | 3 | 2 | 5 | 3 | 4 |
| **O5** | 244 | X | 2 | 5 | 1 | 3 | 3 | 4 |
| Total O5 |  |  | 2 | 5 | 1 | 3 | 3 | 4 |
| **O3** | 1229 |  | 1 | 3 |  | 0 | 1 | 1 |
|  | 274 |  | 1 | 3 |  | 0 | 1 | 1 |
| Total O3 |  |  | 2 | 5 |  | 0 | 2 | 3 |
| **O2** | 244 | X | 1 | 3 |  | 0 | 1 | 1 |
| Total O2 |  |  | 1 | 3 |  | 0 | 1 | 1 |
| **O7** | 671 |  |  | 0 | 1 | 3 | 1 | 1 |
| Total O7 |  |  |  | 0 | 1 | 3 | 1 | 1 |
| **Total** |  |  | **37** | **100** | **40** | **100** | **77** | **100** |

Overall, the panel for genomic characterization was distributed into 18 sequence types (ST). Except for ST111 and ST244, which were associated with O12 and O4 and O2 and O5, respectively, each ST was associated with only one serotype. The most common STs were ST233 (20.8%), associated with serotype O6, ST111 (13.0%), and ST298, associated with O11 (10.4%). The remaining STs were unique or represented between 3.0% and 10.0%. The majority of STs within the P strains were ST233 (27.0%), ST235, and ST111 (13.5% each). The majority of STs of the H strains were ST298 and ST233 (each representing 15.0%) and ST111 (12.5%). This apparent diversity in serotypes and STs was confirmed by the cgMLST results represented in Figure 3. For cgMLST, of a total of 3,867 loci searched, 3,076 loci were present in the genomes of all strains. Overall, strains diverged in distance from 0 to 2,897 loci and were well distributed according to their origin (human/hospital environment), year of collection, carbapenem resistance, and DDAC resistance (Figure 4).

**FIGURE 3.**
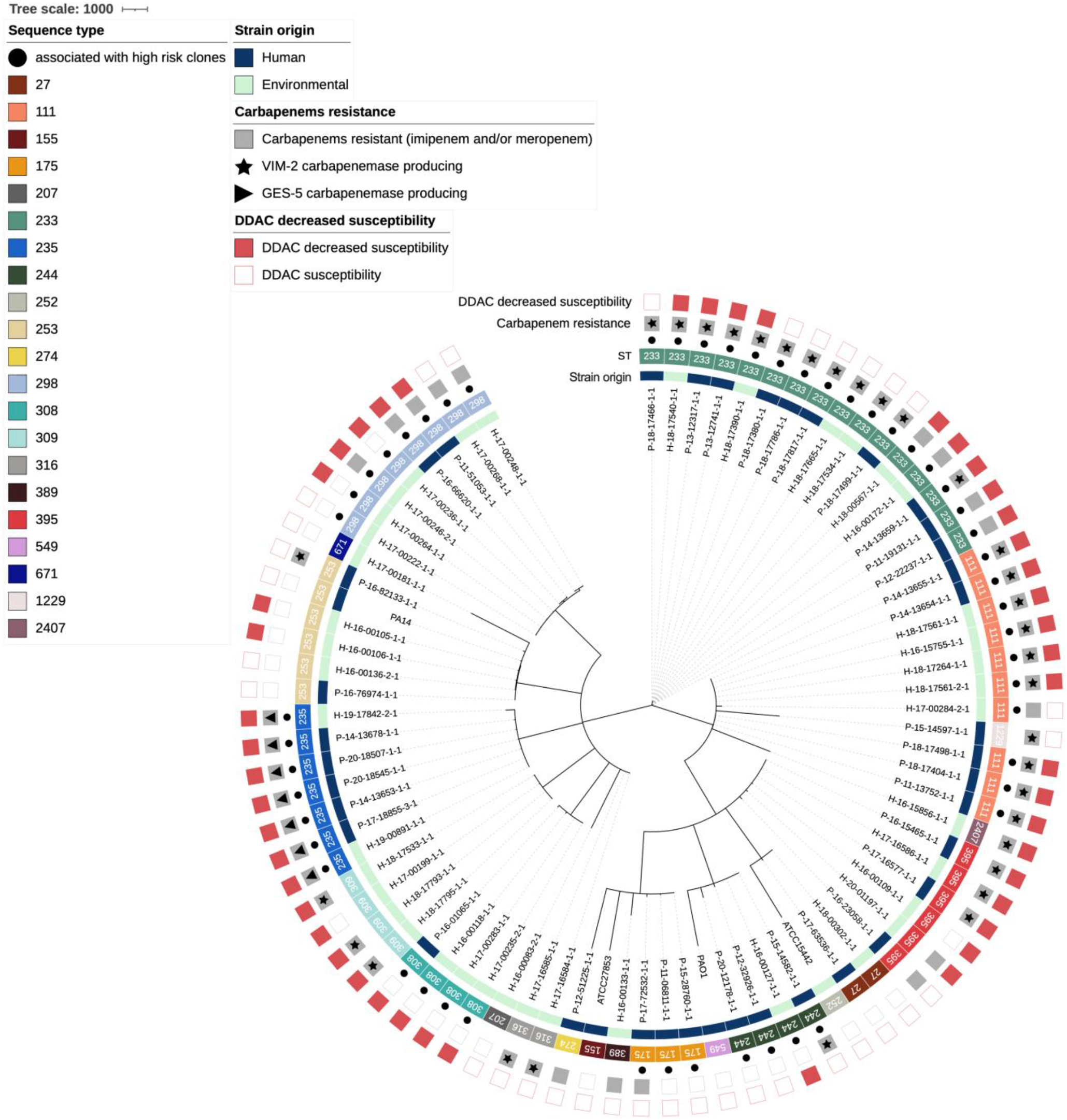
Minimum spanning tree of strains from panel for genomic characterization and reference strains (n=77+4). Core genome MLST clustering according to the cgMLST *Pseudomonas aeruginosa* scheme published by Tönnies *et al*.^[56]^. The reference strains are ATCC15442, ATCC27853, PAO1, and PA14. Were represented: the origin of the strains (human or environmental), sequence type, carbapenem resistance phenotype, whether it was associated with a VIM-2 or GES-5 carbapenemase, and decreased susceptibility phenotype to DDAC. ATCC: American Type Culture Collection; DDAC: didecyldimethylammonium chloride; GES-5: strains producing the class A carbapenemase GES-5; H: strains isolated from the hospital environment; P: strains isolated from patients; ST: Sequence type; VIM-2: strains producing the class B carbapenemase VIM-2.

**FIGURE 4.**
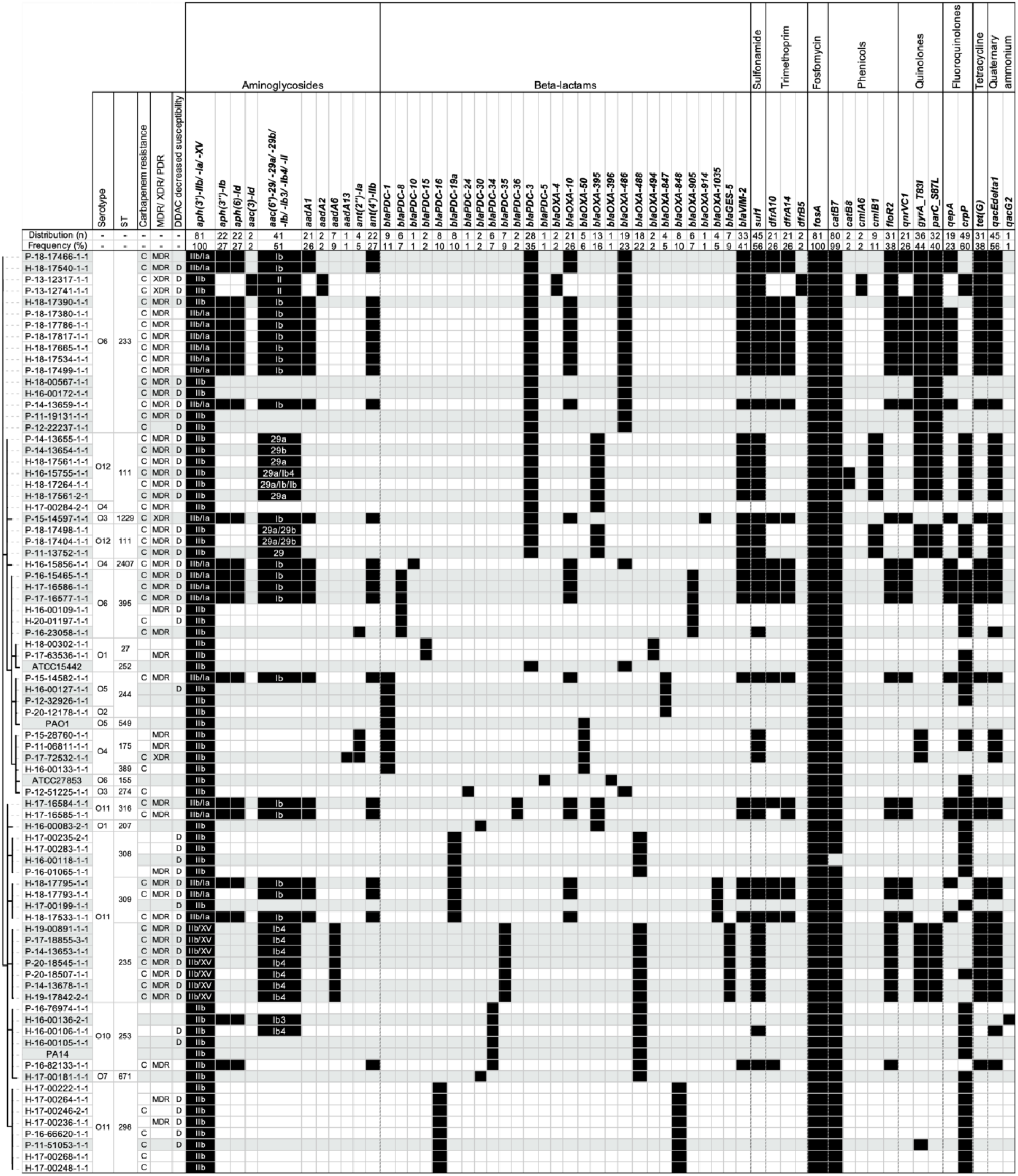
Antimicrobials resistance-associated genes of strains from panel for genomic characterization and reference strains (n=77+4). Strains were organized according to the cgMLST minimum distance tree. The serotype, the sequence type, the presence of a carbapenem resistance (imipenem and/or meropenem) phenotype, whether the strain was multidrug-resistant, extensively drug-resistant, or pandrug-resistant, and the presence of a decreased susceptibility to DDAC phenotype are mentioned. The sub-variants of the *aph(3’)* gene were grouped as well as the sub-variants of *aac(6*’), in case of co-occurrence of these sub-variants they were indicated and separated by a slash. ATCC: American Type Culture Collection; C: strains with carbapenems resistance; D: strains with decreased susceptibility to DDAC; H: strains isolated from the hospital environment; O: Serotype; MDR: multidrug-resistant; P: strains isolated from patients; PDR: pandrug-resistant; ST: Sequence type; XDR: extensively drug-resistant.

Numerous genes conferring antibiotic resistance were found, some constitutive for *P. aeruginosa* and others acquired. At least one resistance gene was associated with each antibiotic class (Figure 4). For aminoglycosides, the resistance genes encoded N-acetyltransferases (*aac* variants), aminoglycoside nucleotidyltransferases (*aad* variants), and aminoglycoside O-phosphotransferases (*aph* variants) were found. Regarding resistance to carbapenems, the genes found comprise the carbapenem-hydrolysing class A β-lactamase *bla*_*GES-5*_ and the subclass B1 metallo-β-lactamase *bla*_*VIM-2*_ (Figure 4). For ß-lactamines, the resistance genes comprise the cephalosporin-hydrolysing class C β-lactamase *bla*_*PDC*_ variants and OXA family oxacillin-hydrolysing class D β-lactamases *bla*_*OXA*_. For sulfonamide, only the dihydropteroate synthase *sul1* was found; for trimethoprim its was found dihydrofolate reductases (*dfrA10, dfrA14*, and *dfrB5*).

For quinolone resistance, genes coding for the pentapeptide repeat protein QnrVC1, the efflux MFS transporter QepA, and modifications of the amino acid sequence in the quinolone resistance determining region (QRDR) due to sequence alterations in position 87 of *parC* and 83 of the *gyrA* genes. Interestingly, the efflux SMR transporters encoding *qacEdelta1* and *qacG2* genes were found but were not statistically associated with the quaternary ammonium resistance (Figure 4).

### Analysis of the overexpression of efflux pumps and cephalosporinase activity

The MIC values of imipenem and meropenem were considerably reduced in the presence of PaβN (in particular at 50 mg/L) for all 13 clinically selected *P. aeruginosa* strains, except for those expressing VIM-2 or GES-5 carbapenemase (SD 1 Part 3). This was particularly noted for those belonging to the ST111 population, for which showed no effect. The cephalosporinase inhibitor cloxacillin showed a much more limited inhibitory effect for all the strains tested. The effect of the inhibitor efflux pump PaβN on response to DDAC was much greater, displaying a ≥ 2-fold decrease in MIC values conferring susceptibility to this quaternary ammonium compound. Interestingly, the cephalosporinase inhibitor cloxacillin had the opposite effect, increasing the DS to DDAC for all strains, including the PA14 and PAO1 reference strains.

Evaluation of the expression of *mexA, mexB, oprM, mexE, mexF*, and *oprN* efflux pumps encoding-genes was performed for three selected strains expressing VIM-2 Compared with PAO1 as control, the two strains that accumulated carbapenem nonsusceptibility and DS to DDAC phenotypes overexpressed *mexA, mexB*, and *oprM* genes (fold-change up to 1.5). For the third strain (nonsusceptible to carbapenem but susceptible to DDAC) *mexE, mexF*, and *oprN* genes were overexpressed. Take all together, these data strongly suggest that the MexA, MexB and/or OprM may have a role in the resistance to DDAC in *P. aeruginosa*, but they are likely to play a role in carbapenem NS phenotype.

## Discussion

Carbapenem-resistant *P. aeruginosa* strains represent an important factor in the prevalence of hospital-acquired infections worldwide. The mechanism of resistance is carried out by various genetic supports, either through the acquisition of resistance genes on mobile genetic elements (i.e., plasmids) or through mutational processes that alter the expression and/or function of chromosomally encoded mechanisms such as efflux pumps and porins. These strategies can severely limit the therapeutic options for the treatment of serious *P. aeruginosa* infections. This ten-year study conducted in a 1,410-bed teaching hospital in Normandy, France evaluated the proportion of resistance to carbapenems (9.5%), which was mainly supported by the acquisition of carbapenem-hydrolyzing enzymes such as VIM-2 carbapenemase (in 39.3% of carbapenem-resistant strains in the study panel). This prevalence is in accordance with numerous hospital studies pointing out that such of isolates were mainly obtained from patients in ICUs^[14]^. Furthermore, the prevalence of MDR/XDR strains is an even more serious therapeutic challenge. The occurrence of these strains was increased in nosocomial infections, particularly for patients with altered health and comorbidities, unfavorable Charlson index score, and/or immunocompromised status. Thus, epidemiological outcome studies have shown that infections caused by drug-resistant *P. aeruginosa* are associated with significant increases in morbidity, mortality, duration of hospital stay, associated chronic care, and the overall cost of treating the infection^[30]^. For treatment efficacy, it is crucial to diagnose *P. aeruginosa* infection/colonization at an early stage in a hospitalized patient. The prescription of the appropriate antibiotic to initiate therapy is essential to optimize the clinical outcome^[31]^. From our clinical data, we reveal that antipseudomonal prescriptions following species identification were present in only 60% of cases and did not correspond to the most active antibiotics such as colistin, amikacin, ceftolozane-tazobactam, aztreonam or meropenem.

Recent studies suggest that *P. aeruginosa* strains have nonclonal population structures with few highly successful clusters^[22]^. This observation was corroborated by our whole genome analyses. Although some genomes had indistinguishable genetic distances, suggesting epidemiologic links, all 77 sequenced strains were well distributed according to their origin (human/environmental), year of collection, or antibiotype. However, among the 18 STs, seven, ST233, ST111, ST298, ST235, ST244, ST308, and ST175, representing 67.5% of the study population. They are considered to be among the top 10 *P. aeruginosa* high-risk clones worldwide based on prevalence, global spread, and association with MDR/XDR profiles and the expressing-carbapenemase strains^[32]^. Thus, carbapenemase-producing isolates were clearly associated with specific clones, such as ST233, ST111, and ST395, and STs represented once only, all of which express VIM-2 enzymes, and with ST235, which carries the GES-5 enzymes. These carbapenemase-producing strains, which are known to be supported by highly transferable plasmids, were expressed more often in MDR and XDR profiles. Their prevalence increased during the study period, suggesting the role of outbreaks and/or a route of dissemination *via* environmental reservoirs, also implying that carbapenemase occurrence in *P. aeruginosa* is primarily due to clonal spread.

One point of interest in our study is the MIC determination of *P. aeruginosa* strains to a quaternary ammonium compound, DDAC, that is widely used in hospital disinfectants and as a biocide in various applications. Considering an MIC>62.9 mg/L, (corresponding to the concentration of DDAC in the commercial disinfectant solution), DS to DDAC existed for 38.9% of strains of the panel and was significantly more associated with MDR and XDR *P. aeruginosa* profiles and more prevalent in the hospital environment (62.5% of them) than in human strains (28.2%). Interestingly, DS to DDAC for P strains trended to be associated in hospital-acquired situation (14.3% in community versus 31.6% in hospital acquisition). Furthermore, the strains showing DS to DDAC circulated throughout the ten-year study period since 2011 and were again estimated at 28.0% by random inclusion among all clinical *P. aeruginosa* strains of our hospital^[33]^. It was previously described that the DDAC MICs of susceptible strains were between 15 mg/L^[34]^ and 20 mg/L^[35]^, which is consistent with our results for a major part of our DDAC-susceptible strains (8-32 mg/L). Interestingly, Goodarzi *et al*.^[36]^ showed that out of 92 *P. aeruginosa* hospital strains isolated between 2019 and 2020 (Hamadan City Hospital, Iran), 32.6% had a DDAC MIC ranging from 64 to 128 mg/L as found in the present study. They also tested the efflux pump inhibitor carbonyl cyanide m-chlorophenyl hydrazone (CCCP) at 10 mg/L, and the results showed that 44.6% of their strains had a two-fold to 64-fold reduction in MIC^[36]^. We also confirmed the important effect of the PaβN inhibitor efflux pump showing a greater decrease in MIC values for DDAC and a lesser decrease for carbapenems (except for carbapenemase-producing strains). This decreased susceptibility may be explained by the intrinsic mechanism of overexpression of the RND efflux pump system such as MexAB-OprM. It is a well-most known drug efflux system constitutively expressed in *P. aeruginosa*, which contributes to intrinsic antibiotic resistance^[37,38]^. Furthermore, Li et al.^[39]^ clearly demonstrated that the cephalosporinase inhibitor cloxacillin is a substrate for this pump. This could explain why cloxacillin showed a much more limited inhibitory effect on carbapenem resistance and even increased the DDAC MICs for all the strains tested. Moreover, we can also confidently exclude a potential role of the efflux SMR transporters *qacEdelta1* and *qacG*, even though they were present in many of our studied isolates.

According to Nasr et al., exposure to subinhibitory concentrations of DDAC leads to reduced susceptibility to amikacin, gentamicin, meropenem, and ciprofloxacin^[24]^. These data reinforce the assumption that antiseptics such as DDAC present in the hospital environment may constitute an important selection pressure leading to the emergence of multidrug resistant strains. Vigilance and monitoring of antimicrobial resistant strains from the close environment of patients appear necessary to prevent hospital-acquired infections. In this context, further studies testing other disinfectants classes as chlorine-based, iodine-based, phenol-based, aldehyde-base disinfectants, alcohol-based, quaternary ammonium-based, and biguanides could be important to lead in the future.

This comprehensive evaluation of the carbapenem resistance mechanisms of *P. aeruginosa* isolates collected over ten years in our hospital highlights the importance of acquired MβL-VIM-2 carbapenemases and chromosomal resistance mediated by the RND efflux pumps system including MexAB-OprM. The assessment of a high proportion of MDR and XDR isolates with DS to a quaternary ammonium compound in our hospital environment is worrisome. This phenomenon was not explained by the dissemination of limited specific strains but appears as the consequence of cumulative mechanisms of resistance regardless of the *P. aeruginosa* population. *P. aeruginosa* strains resistant to molecules that are widely used in medicine and hospital disinfection are probably distributed in hospitals worldwide and can severely limit the therapeutic options for the treatment of serious infections.

## Materials and Methods

### *P. aeruginosa* bacterial strains

#### Reference strains

Four well-described and genome-available reference strains were used in the present study, ATCC27853 and ATCC15442, obtained from the American Type Culture Collection (ATCC), and PAO1 and PA14, from the collection of Institut Pasteur (Paris, France). Strain ATCC15442 is recommended for disinfectant susceptibility testing^[40]^, strain ATCC27853 is the *Pseudomonas* spp. reference for antibiotic susceptibility testing^[41]^, PAO1 is the reference genome for the *P. aeruginosa* species^[42]^ and strain PA14 is a highly virulent isolate representing the most *P. aeruginosa* common clonal group worldwide contrary to PAO1^[43]^.

#### Hospital strains and study panel

Data on all *P. aeruginosa* strains from Caen University Hospital, a 1,410-bed teaching hospital in Normandy, France, were extracted from the laboratory management system (TD-Synergy, Montbonnot-Saint-Martin, France) for human strains (only the first of each patient, screening, or clinical), and from annual reports of the hospital hygiene ward for hospital environment strain^[44]^. In total during the period of 1 January 2011 to 31 December 2020, 13,049 *P. aeruginosa* strains were isolated; 6,661 strains (51.0%) came from patients (P strains, first strain isolated from the patient) and 6,388 (49.0%) from the water hospital environment (H strains). All were stored at room temperature in an agar medium, or at -80 °C on brain-heart infusion medium (BHI; bioMérieux, Marcy-l’Étoile, France) with 15% glycerol (VWR, Radnor, Pennsylvania, USA). Among them, 180 strains were selected retrospectively based on the year and hospital ward isolation and the antibiotype to constitute the “study panel” (Supplementary Data 1 [SD] Part 1 and SD 2). Of these, 124 were from patients (clinical and screening) over the 2011 to 2020 period, and 56 were from the hospital environment over the 2016 to 2020 period (before 2016, H strains could not be included because they were not conserved). All were subcultured on tryptic soy broth (TS; Bio-Rad, Hercules, California, USA) and then stored on 15% glycerinated BHI at -80 °C. The species identification was determined by matrix-assisted laser desorption ionization-time of flight (MALDI-TOF) mass spectrometry (Microflex; Bruker Daltonik, Bremen, Germany). Then, from the study panel, 77 strains were selected based on the resistance profile to represent the variability of antibiotic and disinfectant resistance patterns, the origin of the sample, and the sampling year to constitute the panel for genomic characterization (SD 1 Part 2). Thirty-seven of them were from patients, and the other 40 were from environmental samples. Among them, 13 strains (6 from patients and 7 from the hospital environment, with various resistomes) were selected to determine the overexpression of efflux pumps and cephalosporinase (AmpC), and it constituted the “representative short panel” (SD 1 Part 3).

In parallel to this study, 100 strains of *P. aeruginosa* (10 per year) were randomly selected from the 13,049 strains of Caen UH, to estimate the frequency of nonsusceptibility to DDAC.

#### Strains and patient data (SD 1 Part 1)

Available data were extracted for all 180 strains from the laboratory software (TD-Synergy TECHNIDATA, Montbonnot-Saint-Martin, France), including the year, origin and type of sampling, hospital ward, and carbapenemase-expressing status. In addition, patient information was retrospectively collected, including the patient gender, age, presence of comorbidities, immune status, whether it was a *P. aeruginosa* infection or colonization, whether or not death occurred (*P. aeruginosa*-related or not), and antipseudomonal antibiotic therapy administered during hospitalization (before and after *P. aeruginosa* identification). According to the European Committee on Antimicrobial Susceptibility Testing (EUCAST), the following molecules were considered active against *P. aeruginosa* for medical use: antipseudomonal penicillins (ticarcillin, ticarcillin-clavulanic acid, piperacillin, and piperacillin-tazobactam), carbapenems (doripenem, imipenem, imipenem-relebactam, meropenem, and meropenem-vaborbactam), monobactams (aztreonam), 3^rd^- and 4^th^-generation cephalosporins (cefepime, cefiderocol, ceftazidime, ceftazidime-avibactam, and ceftolozane-tazobactam), aminoglycosides (amikacin, gentamicin, netilmicin, and tobramycin), fluoroquinolones (levofloxacin, ciprofloxacin), fosfomycin and polymyxins (colistin and polymyxin B)^[45]^. The P strain *P. aeruginosa* acquisition type was also determined: community (i.e., the acquisition occurred before or during the first 48 hours of hospitalization) or hospital acquired. The Charlson comorbidity index score was calculated using the available tool at https://www.rdplf.org/calculated^[46]^.

### Antimicrobial susceptibility testing (AST)

#### Antibiotic susceptibility testing

Among the 6,661 P strains, ASTs were available for 4,375 *P. aeruginosa* strains against 16 antipseudomonal molecules categorized into 8 classes. Disk diffusion method was performed for all except for colistin and fosfomycin, tested by broth microdilution by VITEK^®^ 2 (bioMérieux, Marcy-l’Étoile, France) or by the Sensititre™ Vizion™ System (Thermo Fisher Scientific, Waltham, Massachusetts, USA). Interpretation of values according to the guidelines of the Comité de l’antibiogramme de la Société Française de Microbiologie (CASFM) each year, from 2011 to 2020. The data were gathered to estimate the percentage of resistance (R) for each antibiotic and classification in multidrug-resistant (MDR), or extensively drug-resistant (XDR) profiles.

For the 180-strain study panel, antibiotic susceptibility testing was performed again for 16 antipseudomonal antibiotics (Bio-Rad, Hercules, California, USA) in association or not with a β-lactamase inhibitor that are distributed in 7 distinct classes: penicillin (ticarcillin, ticarcillin-clavulanic acid, piperacillin, piperacillin-tazobactam), carbapenems (imipenem, meropenem), monobactams (aztreonam), cephalosporins (cefepime, ceftazidime, ceftolozane-tazobactam), phosphonic acid (fosfomycin), aminoglycosides (amikacin, tobramycin, gentamicin) and fluoroquinolones (ciprofloxacin, levofloxacin). MIC for colistin was not performed. Antibiotic susceptibility testing was performed on Mueller-Hinton agar (Becton Dickinson, Franklin Lakes, New Jersey, USA) following the EUCAST guidelines for *Pseudomonas* spp. disk diffusion method^[41]^. Following the 2021 edition of the EUCAST breakpoint tables for interpretation, breakpoints were set^[45]^. Strains showing resistance in at least three antibiotic classes were considered MDR. Those with remaining susceptibility in one or two antibiotic categories were considered XDR. Pan resistance could be defined as resistance to all eight antibiotic classes^[47]^.

#### Quaternary ammonium susceptibility testing

For the 180-strain study panel, the minimum inhibitory concentration (MIC) of the detergent-disinfectant DDAC was assessed by the reference broth microdilution method using cation-adjusted Mueller-Hinton broth w/TES (Thermo Fisher Scientific, Waltham, Massachusetts, USA) at concentrations ranging from 0.5 to 1024 mg/L in triplicate per strain. The threshold of decreased susceptibility (DS) in the present study was MIC>62.9 mg/L, corresponding to the concentration of DDAC in the disinfectant solution according to the manufacturer’s instructions.

Furthermore, for the 100 randomly selected *P. aeruginosa* strains between 2011 to 2020 the MIC of DDAC was determined under the same technical conditions^[33]^.

### Whole-genome sequencing and bioinformatic analysis

For the reference strains, ATCC27853 and PA14 genomes were obtained from the European Nucleotide Archive (ENA) database with accession numbers CP015117 and ASWV01000001, respectively, while the ATCC15442 and PAO1 sequences were obtained from GenBank with accession numbers GCF_000504485.1 and GCA_000006765.1, respectively.

The 77 strains were sequenced by the “Plateforme de Microbiologie Mutualisée P2M” (Institut Pasteur, Paris, France). A MagNA Pure 96 instrument (Roche Diagnostics, Meylan, France) was used for DNA extraction, a Nextera XT library kit (Illumina Inc., San Diego, USA) was used for NGS library construction, and a NextSeq500 (Illumina Inc., San Diego, USA) was used for sequencing. FastQC V0.11.0^[48]^ and MultiQC V1.9^[49]^ software were used for quality control checks on raw sequence data. The paired-end reads were preprocessed (filtered and trimmed) using fqCleanER (https://gitlab.pasteur.fr/GIPhy/fqCleanER), with a minimal read size of 70 bp and a Phred quality score of 28. De novo assembly was performed using SPAdes 3.12^[50]^ with a 50X minimum average sequencing depth, and Quast software V5.0^[51]^ was used for final assembly quality checks.

Species identifications based on the sequences have been validated with Ribosomal Multilocus Sequence Typing (rMLST), available at https://pubmlst.org/^[52]^. *In silico, P. aeruginosa* strain serotyping was performed using PAst1.0 software, available at https://cge.cbs.dtu.dk/services/PAst-1.0/^[53,54]^. Then, multilocus sequence typing (MLST) was performed on the sequence variation of 7 housekeeping genes using MLST2.0 software (version 2.0.4, 2019/05/08; database version 2021/10/04). This software uses the MLST allele sequence and profile obtained from PubMLST.org^[53,55]^. Finally, the core genome MLST was determined based on the *P. aeruginosa* MLST scheme targeting 3,867 loci (available on cgmlst.org at https://www.cgmlst.org/ncs/schema/16115339/locus/, database version 2021/05/26)^[56]^. Chewbacca software version 2.8.5^[57]^ was used for the cgMLST scheme conversion and allele calling. Finally, a neighbor-joining tree based on cgMLST was visualized with iTOL v6.3.3^[58]^. The nucleotide sequences were submitted to AMRFinderPlus analysis (version 3.10.30, database version 2022-05-26.1)^[59]^ with a minimal identity of 80% and minimal coverage of 50% to identify antimicrobial resistance genes and known resistance-associated point mutations. All 77 assembled genomes were deposited in the BioProject PRJNA884650.

### Analysis of the overexpression of efflux pumps and cephalosporinase activity

Imipenem, meropenem, and DDAC MICs values were determined in the presence/absence of the efflux pump inhibitor phenyl-arginine-β-naphthylamide (PaβN at 25, 50 and 100 mg/L) and the cephalosporinase (AmpC) inhibitor cloxacillin (at 250 mg/L). Susceptibility testing was performed by broth microdilution method as previously described for the 13 strains of the representative short panel and 2 reference strains (PAO1 and PA14). In addition, the expression of the *mexA, mexB, oprM, mexE, mexF*, and *oprN* genes was determined by quantitative real-time PCR. Total RNA was extracted from bacterial cells to the late exponential phase using the Direct-Zol RNA miniprep kit (Zymo Research, Irvine, California, USA), and the residual chromosomal DNA was removed using a Turbo DNA-free kit (Life Technologies, Carlsbad, California, USA). A NanoDrop One spectrophotometer (Thermo Fisher Scientific, Waltham, Massachusetts, USA) was used for quantification, and cDNA was synthesized from total RNA using a QuantiTect reverse transcription kit (Qiagen, Hilden, Germany) following the manufacturer’s guidelines. The transcript levels were determined by the ΔΔCT method (CT is the threshold cycle) using the expression of the housekeeping *gyrB* gene as a reference transcript. The level of transcription measured for the PAO1 reference strain was used to determine the expression ratios for the strains of interest. The sequences of the primers associated with each gene are listed in SD 3.

### Statistical analysis

All statistical tests were performed with GraphPad Prism version 9.0.0 for macOS (GraphPad Software, San Diego, California USA). Independence tests of populations using Fisher’s exact test were performed successively to determine if the DS to DDAC was linked to nonsusceptibility of strains (MDR or XDR phenotypes) and/or if the DS to DDAC was linked to carbapenem resistance. A chi-square test was performed to compare the distribution of DS to DDAC according to the strain origin. Carbapenem resistance average annual expression frequency was obtained by averaging annual expression frequency obtained for each year between 2011 and 2020, and 95% confidence intervals were calculated by the Wilson/Brown method.

## Data Availability

All data produced in the present study are available upon reasonable request to the authors

## Data availability statement

All data generated or analyzed during this study are included in this published article (and its Supplementary Information files).

## Acknowledgements

The authors wish to thank Dr. Charles Maurille for his help in determining the comorbidities and immune status of patients. Part of this work was performed on computing resources provided by CRIANN (Normandy, France, https://www.criann.fr/).

## Author Contributions

Conceptualization, M.P., F.Gr., A.L. and S.L.H; methodology, M.P., S.C., F.Gu., M.A., B.L., and F.Gr.; software, M.P. and F.Gr.; validation, F.Gr., A.L. and S.L.H.; formal analysis, M.P. and F.Gr.; investigation, M.P. and S.L.H.; data curation, M.P., A.L., F.Gr. and S.L.H; writing—original draft preparation, M.P.; writing—review and editing, M.P., S.C., F.Gr, F.Gu., B.L, M.A, JC.G., S.L.H. and A.L.; visualization, M.P., S.C., F.Gr., B.L., JC.G., A.L. and S.L.H.; supervision, A.L. and S.L.H.; project administration, A.L. and S.L.H.; funding acquisition, A.L. and S.L.H. All authors have read and agreed to the published version of the manuscript.

## Additional information

### Ethics Statement

This study has been conducted in compliance with the Helsinki Declaration (ethical principles for medical research involving human subjects) and in accordance with the guidelines of research board of our teaching hospital, Caen, France. Ethic committee of CHU Caen Normandie reviewed and approved the study number ID 3784. It was a non-interventional study: specimens used in this study were part of the routine patient management without any additional sampling. No objection has been provided by patients or their families.

### Funding

This work was financially supported by Normandy Regional Council and French Ministry of Agriculture (EcoAntibio Plan).

### Conflict of Interest

The authors declare that the research was conducted in the absence of any commercial or financial relationships that could be construed as a potential conflict of interest.

## Supplementary Material

The Supplementary Material for this article can be found online.

## Notes

### Competing Interest Statement

The authors have declared no competing interest.

### Author Declarations

Ethic committee of CHU Caen Normandie reviewed and approved the study number ID 3784

