## supplemental materials for "A ten-year microbiological study of *Pseudomonas aeruginosa* strains revealed diffusion of carbapenems and quaternary ammonium compounds resistant populations"

### *Supplementary Material*

**Supplementary Data 1. Characteristic of the strains used in the study panel (Part 1), in the panel for genomic characterization (Part 2) and in the representative short panel (Part 3).**

Please see the joined Excel file named Supplementary_data1.xls

**Supplementary Data 2. Selection diagram for the study and reference strains.** [1]: 6 classes tested, i.e., 16 antibiotics for hospital use; [2]: didecyldimethylammonium chloride; ATCC: American Type Culture Collection; H: strains isolated from the hospital environment; MALDI-TOF: Matrix Assisted Laser Desorption Ionisation/Time Of Flight; P: strains isolated from patients; PSAE: *Pseudomonas aeruginosa*; UHC: University Hospital Center; WGS: Whole Genome Sequencing.

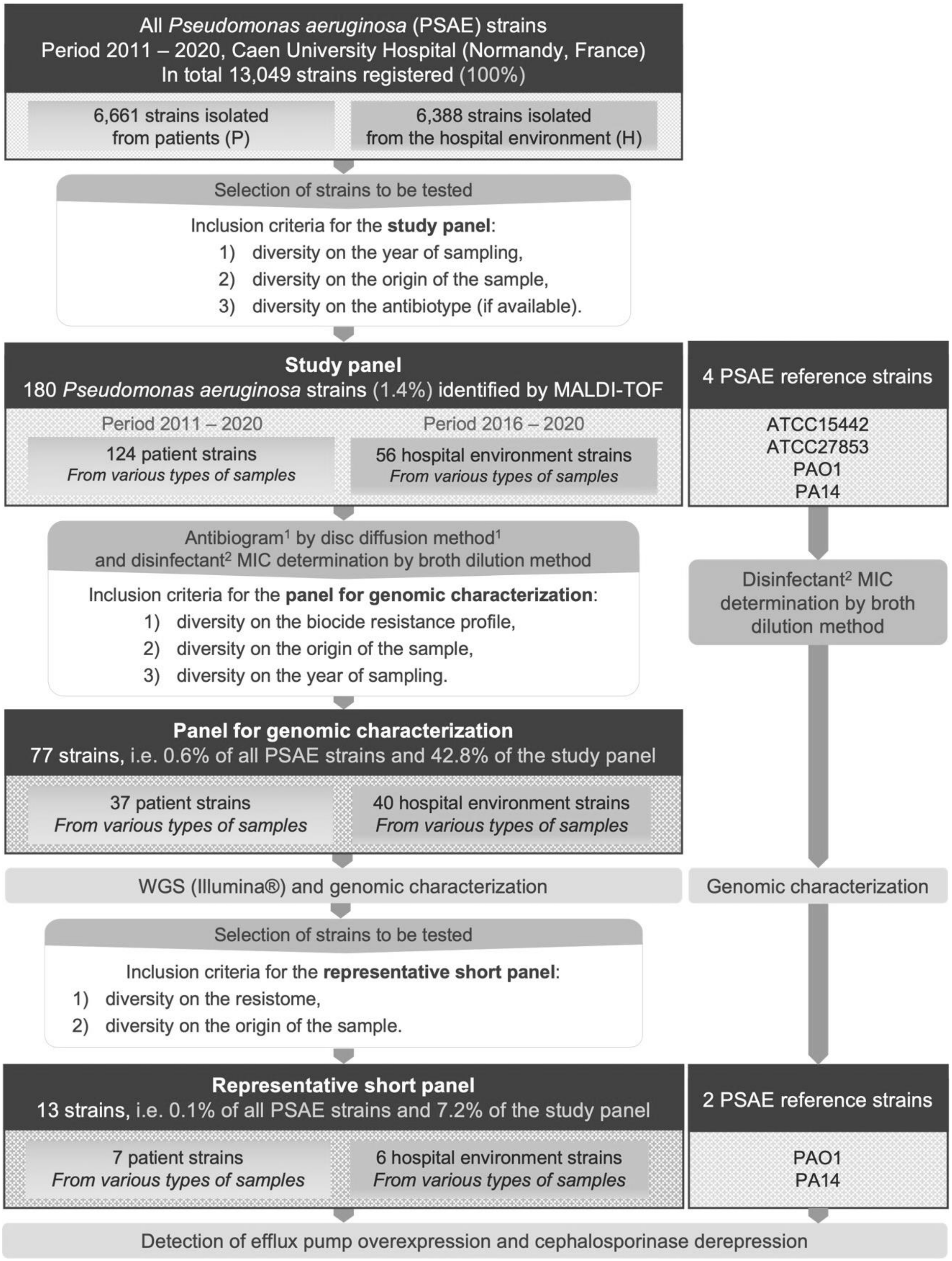

**Supplementary Data 3. The sequence of oligonucleotide primers.**

| Target gene | Type | Sequence 5'-3' |
|---|---|---|
| *mexA* | Forward | CTGGAAGGTCGCCTCGAATT |
| | Reverse | AGGATGGCCTTCTGCTTGAC |
| *mexB* | Forward | CGATCGTGATGACCTCCCTG |
| | Reverse | TACCCAGAAGATCGCCAGGA |
| *oprM* | Forward | CCTGGACTACGCGAAGATCC |
| | Reverse | GAGCTGGTAGTACTCGTCGC |
| *mexE* | Forward | GGTCTACGCCTACTTCGACG |
| | Reverse | CCTGGTTGTCGAGGAAGTCC |
| *mexF* | Forward | TGATCCTGATCGTGCCGATG |
| | Reverse | GCGAACTCGACGATCAGGAT |
| *oprN* | Forward | GGGTCTGTTCAGTCTGCTG |
| | Reverse | GGTCGGATCGTCGAACTGTT |
| *gyrB* | Forward | CACGTACGACTCTTCCAGCA |
| | Reverse | GAACACCATGTGGTGCAGAC |

**Supplementary Data 4. Temporal distribution: of all *Pseudomonas aeruginosa* strains isolated at Caen UHC over 2011-2020, the study panel, and of genomic characterization panel.**

| | | **All *P. aeruginosa* strains isolated (N=13,049)** | | **Study panel (n=180)** | **Panel for genomic characterization (n=77)** |
|---|---|---|---|---|---|
| | Sampling year | Strains/year | **Average strains/year** [IC95%] | Strains/year | Strains/year |
| P-strains | **2011** | 630 | **666.1** [615.1-717.1] | 11 | 4 |
| | **2012** | 526 | | 14 | 3 |
| | **2013** | 576 | | 12 | 2 |
| | **2014** | 655 | | 14 | 5 |
| | **2015** | 659 | | 12 | 3 |
| | **2016** | 737 | | 12 | 6 |
| | **2017** | 715 | | 11 | 4 |
| | **2018** | 725 | | 16 | 7 |
| | **2019** | 722 | | 10 | - |
| | **2020** | 716 | | 12 | 3 |
| | **Total** | 6,661 | - | 124 | 37 |
| H-strains | **2011** | 283 | **638.8** [389.2-888.4] | - | - |
| | **2012** | 284 | | - | - |
| | **2013** | 291 | | - | - |
| | **2014** | 310 | | - | - |
| | **2015** | 589 | | - | - |
| | **2016** | 1,026 | | 20 | 11 |
| | **2017** | 542 | | 19 | 14 |
| | **2018** | 989 | | 12 | 12 |
| | **2019** | 957 | | 4 | 2 |
| | **2020** | 1,117 | | 1 | 1 |
| | **Total** | 6,388 | - | 56 | 40 |
| **General total** | | 13,049 | **1,304.9** [1,012.1-1,597.7] | 180 | 77 |

**Supplementary Data 5. Distribution by sample type of the study panel (n=180).** H: strains isolated from the hospital environment; P: strains isolated from patients.

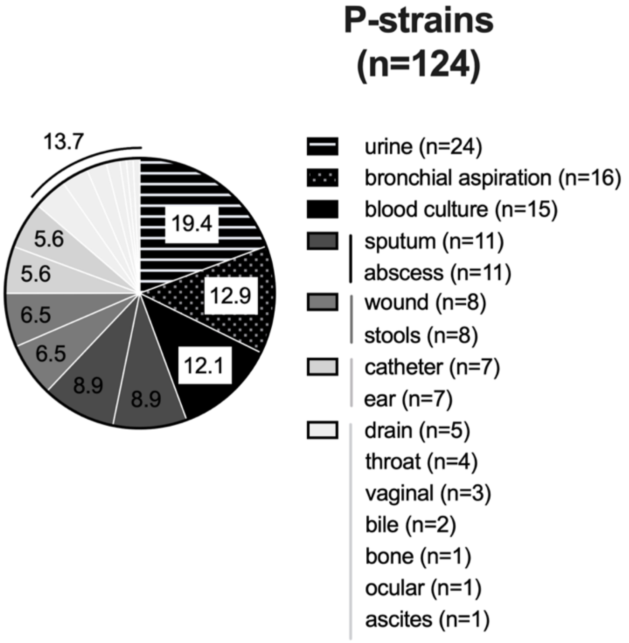


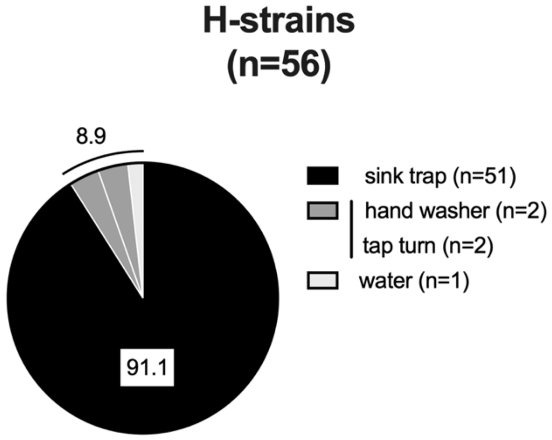

**Supplementary Data 6. Antimicrobial resistance for the study panel (n=180).** (**a**) Occurrence of antibiotics resistance for patient strains (n=124) and hospital environment strains (n=56) of the study panel. For antibiotics, resistance breakpoints were set according to the CASFM/EUCAST 2021, or 2019 edition if a breakpoint was not available, and for DDAC, the DS breakpoint was the concentration in the disinfectant solution following the manufacturer's instructions. Fisher’s test for populations independence for human and  environmental hospital strains resistance frequency. (**b**) Number of strains (in percent) of the study panel (n=180) showing resistance for 0 to 7 antibiotic classes. The hatched areas represent profiles that also accumulate DS to the disinfectant DDAC. The different antibiotic categories are penicillins (ticarcillin, ticarcillin-clavulanic acid, piperacillin, piperacillin-tazobactam), carbapenems (imipenem, meropenem), monobactam (aztreonam), cephalosporins (ceftazidime, ceftolozane-tazobactam, cefepime), phosphonic acid (fosfomycin), aminoglycosides (amikacin, tobramycin, gentamicin) and fluoroquinolones (ciprofloxacin and levofloxacin) were considered. Fisher’s test for populations independence for DS to DDAC and loss of susceptibility to more than three categories of antibiotics. ns: p-value > 0.05, *: p-value ≤ 0.05, **: p-value ≤ 0.01, and ****: p-value ≤ 0.0001 after analysis by Fisher’s independence test. DDAC: didecyldimethylammonium chloride; DS: decreased susceptibility; H: hospital environment;  MDR: multidrug-resistant; P: strains isolated from patients; XDR: extensively drug-resistant.

**a**

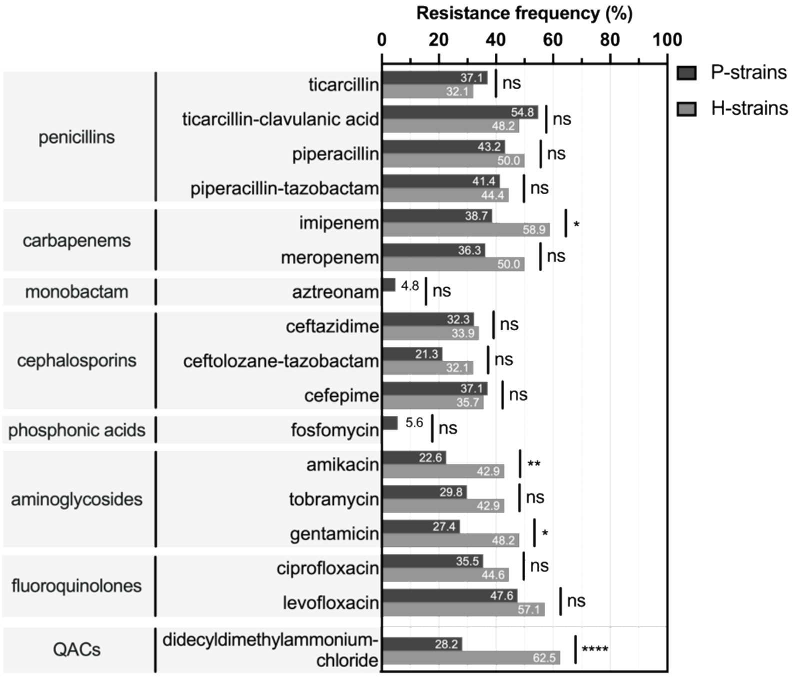


**b**

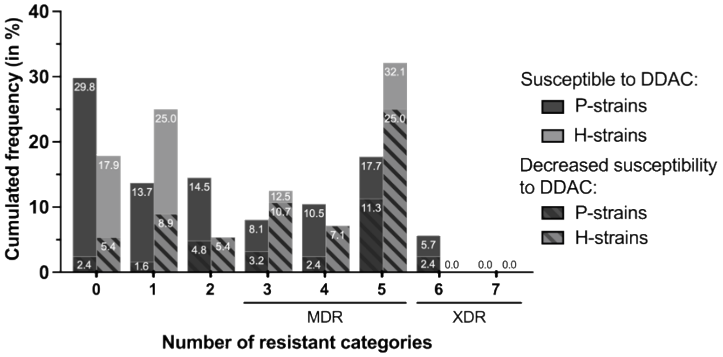
